# Impact of non-pharmaceutical interventions against COVID-19 in Europe: A quasi-experimental study

**DOI:** 10.1101/2020.05.01.20088260

**Authors:** Paul R Hunter, Felipe J Colón-González, Julii Brainard, Steven Rushton

**Author notes:** **DECLARATIONS**. **Data:** The datasets used are in the public domain with copies available from the authors. **Author contributions** PRH and JB conceived of the study. PRH and JB collected the data. PRH, FCG and SR undertook and refined analysis. PRH wrote the first draft which was revised by all authors. Many sharp-eyed readers of the preprint contributed many small corrections via.

## Abstract

The current epidemic of COVID-19 is unparalleled in recent history as are the social distancing interventions that have led to a significant halt on the economic and social life of so many countries. However, there is very little empirical evidence about which social distancing measures have the most impact. We report a quasi-experimental study of the impact of various interventions for control of the outbreak. Data on case numbers and deaths were taken from the daily published figures by the European Centre for Disease Control and dates of initiation of various control strategies from the Institute of Health Metrics and Evaluation website and published sources. Our complementary analyses were modelled in R using Bayesian generalised additive mixed models (GAMM) and in Stata using multi-level mixed effects regression models. From both sets of modelling, we found that closure of education facilities, prohibiting mass gatherings and closure of some non-essential businesses were associated with reduced incidence whereas stay at home orders and closure of all non-businesses was not associated with any independent additional impact. Our results could help inform strategies for coming out of lockdown.

## INTRODUCTION

The current pandemic of COVID-19 is unprecedented in recent history. Not only is the impact of the epidemic being measured by the number of cases and deaths, but also by its impact on overloaded health services and deleterious impacts on quality of life and near-future economic prospects. Wider society was subjected to an almost total stasis of social and cultural life. The benefits of social distancing was shown earliest in China, Italy and Spain that turned the tide on their country’s epidemics using often severe social distancing strategies. What these examples do not do is indicate the relative importance of the different non-pharmaceutical/ social distancing interventions. Given the potentially high economic and social costs arising from stringent control measures (1-5), it has been imperative to determine which social distancing measures are most effective at controlling the pandemic. Imposition and relaxation of control measures should be informed by such knowledge. Early on in pandemic response, much policy was driven by the results of mathematical models (6). However, there has been debate about the validity and limitations of the different models for policy making and modelling approaches that have been used (7-10). It is also useful to assess empirical evidence of what aspects of currently applied non-pharmaceutical control measures have or have not been effective.

A quasi-experimental study design is an observational study where the allocation to receive the intervention (or not) is not randomly made (11). Most European states introduced a similar suite of interventions aimed at reducing contact between individuals to reduce transmission. However, the different types of intervention used and their timing varied from one country to another. No measure was imposed by all European countries. Where measures were put in place, they were often imposed at different points in the development of the epidemics. By late April 2020, some European countries were easing control measures so late April was a good point to take stock of intervention effects. This situation offered a unique opportunity to investigate the putative impacts of the various types of intervention, as each individual-country epidemic forms what is effectively a chrono-sequence of disease spread. The intervention strategies could then be compared as interrupted time series.

We report here analyses of trends in both reported cases and deaths across 30 European countries with rather different approaches to and timing of restrictions. We use a quasi-experimental approach to identify what affects such restrictions may have had on the control of the epidemic.

## METHODS

### Data

Data on new cases and deaths reported by all countries were obtained by the European Centre for Disease Control (https://www.ecdc.europa.eu/en/publications-data/download-todays-data-geographic-distribution-covid-19-cases-worldwide). Data up to 24^th^ April are included. For the UK we used only pillar 1 case numbers. Pillar 1: swab testing in Public Health England laboratories and National Health Service hospitals for those with a clinical need, and health and care workers. Pillar 2 results (swab testing for health, social care and other essential workers and their households) as reported daily on https://www.gov.uk/guidance/coronavirus-covid-19-information-for-the-public#history were removed from the case numbers, as pillar 2 sampling was only introduced late in the course of the UK epidemic and inflated total case numbers relative to earlier in the UK outbreak. We also adjusted by the number of tests reported per 1 million population taken as of 16^th^ April from worldometer (https://www.worldometers.info/coronavirus/). In order to compare time series for different countries with different dates of onset for their own epidemics we chose to define the onset as the first day after the latest time where there were two or more consecutive days with no cases reported.

The dates when (if at all) each of the various social restrictions were imposed for 30 European countries were given by the Institute of Health Metrics and Evaluation Data (IHME) (https://covid19.healthdata.org/). The six categories are “Mass gathering restrictions”, “Initial business closure”, “Educational facilities closed”, “Non-essential services closed” and “Stay at home order” and “Travel severely limited”. However, no country was listed in the dataset as having severe travel restrictions and this was dropped from any further analysis. The IHME definitions of these measures are given on their website. Paraphrasing the definitions.

- Mass gathering restrictions are mandatory restrictions on private or public gatherings of any number.
- The first time that there was any mandatory closure of businesses, not necessarily all businesses. Usually such initial closures would primarily affect business such as entertainment venues, bars and restaurants.
- Where non-essential businesses are ordered to close, this usually include many more businesses than were in the first closure category. The second wave of closures likely includes general retail stores and services such as hairdressers.
- Education facilities closure includes all levels (primary, secondary and higher) education that stop direct teacher to student teaching.
- Stay at home orders affect all individuals unless travelling for essential services. They allow close contact only with people of the same household and perhaps some outdoors exercise.

All models adjusted for when countries started to advise or mandate their citizens to wear face masks or coverings (dates of face cover measures are listed in Supplemental Material 1). We do not assess face cover wearing as an independent control measure because of heterogeneity in how the wearing of face coverings in the community was encouraged or mandated and in what contexts. We also adjusted by the number of tests reported per 1 million population taken on the 17^th^ April from WorldoMeter (https://www.worldometers.info/coronavirus/).

### Analyses

We undertook two sets of analyses.

The first analysis was done in R using Bayesian generalised additive mixed models (GAMM) to adjust for spatial dependency in disease between nation states. The variance in the COVID-19 data was four orders of magnitude larger than the mean number of cases, and three orders of magnitude larger than the mean number of deaths. Consequently, models were fit using a negative binomial specification to account for potential over-dispersion in the data.

Let *Yi,t* be the number of COVID-19 cases or deaths for country *i* = 1, ⋯, *I* at time *t* = 1, ⋯, *T* The general algebraic definition of the models is given by:

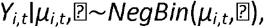

where *Yi,t* is the number of COVID-19 cases or deaths for country *i* = 1, ⋯, *I* at time *t* = 1, ⋯, *μ*_*i,t*_ is the predicted number of COVID-19 cases or deaths for country *i* and time *t*, and ⍰ > 0 is the negative binomial dispersion parameter. A logarithmic link function of the expected number of cases or deaths was modelled as:

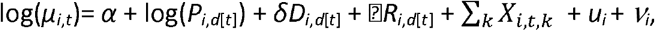

where *α* corresponded to the intercept; log(*P*_*i,d*[*t*]_) denotes the logarithm of the population at risk for country *i* and day *d*[*t*] was included as an offset to adjust case counts by population. *D*_*i,d*[*t*]_ is a linear term for the number of days since the outbreak started, with coefficient *δ. R*_*i,d*[*t*]_ is a linear function of the number of COVID-19 tests carried out per country *i* at day *d*[*t*], with regression coefficient ⍰. *X* is a matrix of *k* intervention measures (e.g. school and business closures) with regression coefficients *β*. Intervention measures comprise of an index of 1, ⋯,*N* number of days following the intervention being implemented. We assumed that the imposition of each intervention led to cumulative changes in effect. Intervention measures were included in the model as a random effect to account for potential non-linearities in the exposure-response relationship. Unknown confounding factors with spatial dependency that represent, for example, human mobility, were incorporated using spatially correlated (i.e. structured) random effects (*u*_*i*_) and independent, identical and normal distributed (i.e. unstructured) random effects (*ν*_*i*_) for each country *i*. Spatial random effects were specified using a Besag-York-Mollie model to account for spatial dependencies and unstructured variation between countries (12). Goodness of fit was evaluated using the Deviance Information Criterion (DIC). Models were fitted in R version 3.6.1 using the INLA package.

The second analysis was a multi-level mixed effects regression analysis in STATA v 16.1. We used a mixed effects negative binomial regression model with cases or deaths on a specific day as the outcome variable, country population as the exposure variable, country as a mixed effect, and days from start of the epidemic as a fixed effect. All main interventions were included as categorical fixed effects in the model as the number of weeks after the start of the intervention with day 1 being the day following the intervention implementation. In sensitivity and collinearity checks (using Stata), we dropped each of the main predictor variables (intervention timings) from the final equation and noted if the regression parameter and standard errors of remaining predictor variables changed dramatically or if the coefficients reversed trend (eg., went from suggesting increase to suggesting decrease).

We also checked for collinearity between the predictor variables by calculating the variance inflation factors (VIF) for the predictors and by calculating the condition number using the coldiag2 command in STATA. A VIF of < 10 suggests that predictor data do not have multi-collinearity problems. VIF values > 10.0 need to be considered with regard to other model diagnostics, such as condition index and eigenvalues. A condition number > 15 with any variance proportions above 0.9, or if eigenvalues were < 0.01 could suggest collinearity that undermines confidence in coefficient estimates, according to guidance in Chatterjee and Hadi 2015 (13) and Regorz 2020 (14). In addition, as sensitivity analysis, within analysis 2 we reran the model dropping each predictor variable in turn to determine whether or not the regression parameters and their standard errors were changed substantially.

## RESULTS

Table 1 gives the estimated date of the start of the epidemic in each country and when each of the five intervention types were implemented, according to the IHME website. “Travel severely limited” was not introduced in any European country. “Mass gathering restrictions”, “initial business closure”, “educational facilities closed”, “non-essential services closed” and “stay at home order” were respectively implemented by 29, 28, 29, 23 and 19 countries. In three countries (Germany, Italy and Spain) the restrictions were not implemented uniformly through the country and so we took the median date. Italy was the first country to enter the epidemic on 22^nd^ February and Lithuania the last on 14^th^ march. Half of all countries had their epidemic start on or before 27^th^ February.

**Table 1.** Timing of estimated start of each country’s main epidemic and the introduction of social distancing measure across 20 European countries (all dates in 2020).

| Country | Start of main epidemic | Mass gathering restrictions | Initial business closure | Educational facilities closed | Non-essential services closed | Stay at home order | Face covering encouraged or compulsory |
| --- | --- | --- | --- | --- | --- | --- | --- |
| Austria | 26/02 | 10/03 | 16/03 | 16/03 | 16/03 | 16/03 | 06/04 |
| Belgium | 02/03 | 13/03 | 13/03 | 14/03 | 18/03 | 18/03 |  |
| Bulgaria | 12/03 | 13/03 | 13/03 | 13/03 | 13/03 | 17/03 | 30/03 |
| Croatia | 11/03 | 09/03 | 19/03 | 16/03 | 19/03 | 17/03 |  |
| Rep of Cyprus | 10/03 | 24/03 | 24/03 | 13/03 | 24/03 | 24/03 |  |
| Czech Rep | 02/03 | 10/03 | 10/03 | 10/03 | 14/03 | 16/03 | 18/03 |
| Denmark | 27/02 | 18/03 | 18/03 | 16/03 |  |  |  |
| Estonia | 11/03 | 13/03 | 13/03 | 16/03 |  |  | 05/04 |
| Finland | 27/02 | 12/03 | 18/03 | 18/03 | 04/04 |  |  |
| France | 26/02 | 04/03 | 14/03 | 12/03 | 14/03 | 16/03 | 05/04 |
| Germany | 26/02 | 22/03 | 17/03 | 16/03 | 23/03 | 22/03 | 01/04 |
| Greece | 05/03 | 08/03 | 12/03 | 11/03 | 22/03 | 23/03 |  |
| Hungary | 05/03 | 12/03 | 12/03 | 16/03 | 16/03 | 28/03 |  |
| Ireland | 04/03 | 12/03 | 15/03 | 12/03 | 24/03 | 27/03 |  |
| Italy | 22/02 | 11/03 | 11/02 | 05/03 | 11/03 | 11/03 | 06/04 |
| Latvia | 08/03 | 13/03 |  | 12/03 |  |  |  |
| Lithuania | 14/03 | 15/03 | 14/03 | 16/03 | 15/03 | 15/03 | 01/04 |
| Luxembourg | 07/03 | 13/03 | 18/03 | 16/03 | 18/03 |  | 20/04 |
| Malta | 08/03 |  | 17/03 | 13/03 | 23/03 |  |  |
| Netherlands | 28/02 | 10/03 | 21/03 | 15/03 |  |  |  |
| Norway | 27/02 | 12/03 | 12/03 | 12/03 |  |  | 05/04 |
| Poland | 07/03 | 10/03 | 31/03 | 12/03 |  | 24/03 |  |
| Portugal | 03/03 | 19/03 | 16/03 | 16/03 | 19/03 | 19/03 | 16/04 |
| Romania | 04/03 | 06/03 | 21/03 | 11/03 | 21/03 | 23/03 |  |
| Slovakia | 07/03 | 12/03 | 16/03 | 12/03 | 16/03 |  | 14/03 |
| Slovenia | 05/03 | 12/03 | 15/03 | 16/03 | 15/03 | 20/03 | 29/03 |
| Spain | 25/02 | 15/03 | 15/03 | 14/03 | 15/03 | 15/03 | 13/04 |
| Sweden | 27/02 | 11/03 |  |  |  |  |  |
| Switzer'd | 26/02 | 28/02 | 16/03 | 13/03 | 16/03 |  |  |
| UK | 28/02 | 23/03 | 20/03 | 23/03 | 24/03 | 23/03 |  |

### Analysis 1

Model metrics are presented in Table 2, while the model results (effects of each intervention on cases and deaths) are shown in Table 3. The dispersion parameter evaluates whether the model is able to cope with potential dispersion in the data. When the value is close to 1 (as it is here) the model is shown to do well at accounting for dispersion. The coefficients on case detection and deaths do not go below the reference of 1.0 (pre implementation value) until 14 days for most interventions, especially mass gatherings and school closure measures. Apparent increases in the first 10 or days after interventions were implemented for cases or deaths reflect expected time for disease development and progression after exposure. The increase is very likely to continue for the first 10-14 days because of exponential disease spread that happened before measures were imposed. This pattern fits with the understood disease incubation, development and concurrent ascertainment processes. The median incubation period is understood to be 4-7 days (15-17), while case ascertainment tended to require an elapse of 2-10 more days (18). For severe cases (those who are hospitalised), 8-14 days post symptom onset tends to coincide with start of a 5-7 day long period of peak disease severity (19). As a result, we expect no intervention could have strongly affected case counts in under about 7 days, and no intervention could strongly reduce counts of death in less than 2-3 weeks.

**Table 2.** Model metrics

| Model | Deviance Information<br>Criterion | Watanabe-Akaike<br>Information Criterion | Conditional predictive<br>ordinate | Dispersion |
| --- | --- | --- | --- | --- |
| Cases | 18009.4 | 18012.6 | -9006.6 | 1.01 |
| Deaths | 8032.4 | 8035.9 | -4018.4 | 0.89 |

**Notes:** The Watanabe-Akaike Information Criterion is described by Watanabe 2010 (44) and was developed to specifically help identify best model fit in Bayesian models. Smaller W-AIC values mean better fit compared to alternative model specifications. The conditional predictive ordinate is a Bayesian diagnostic (45) that detects surprising observations.

**Table 3.**
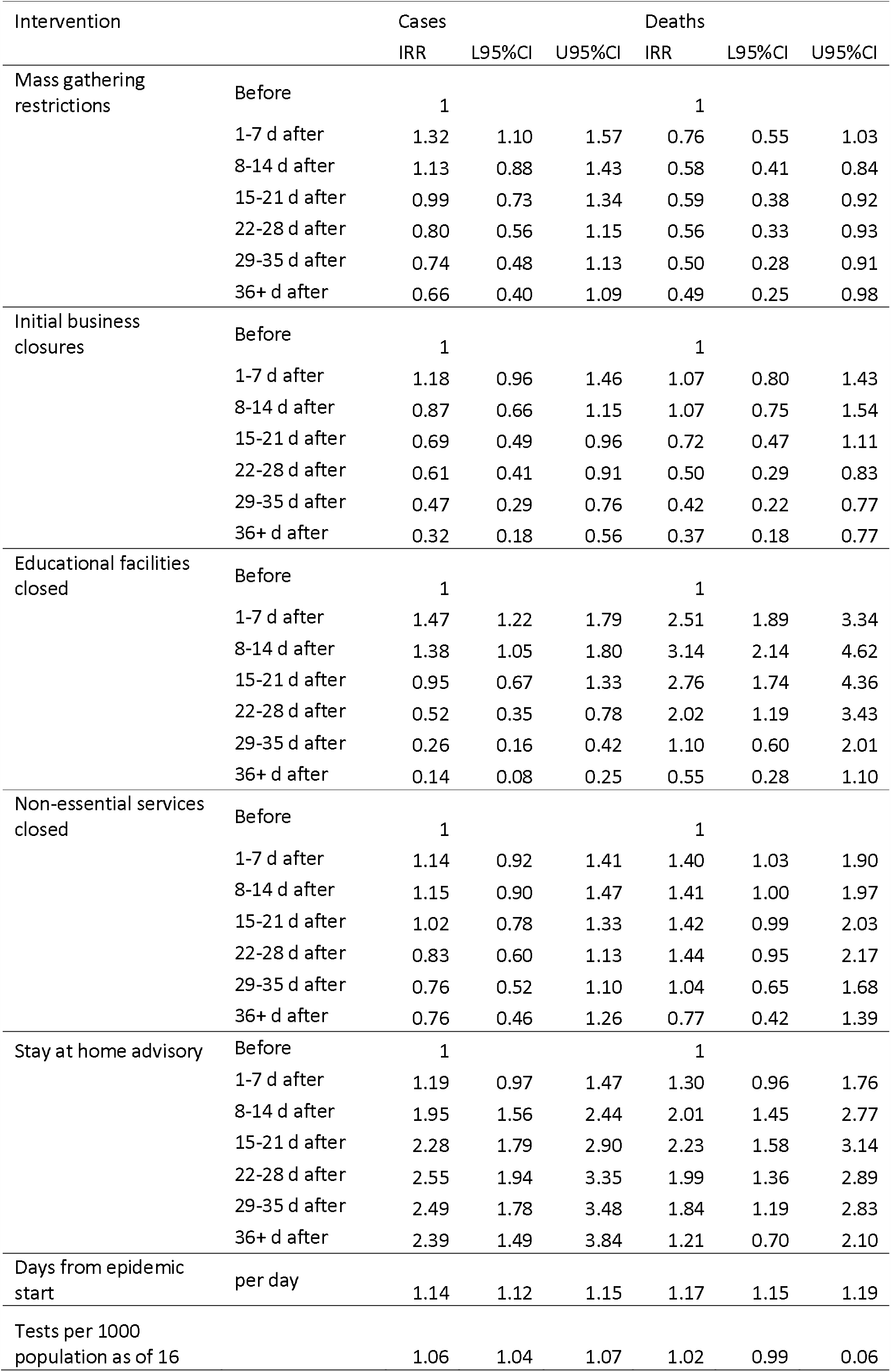
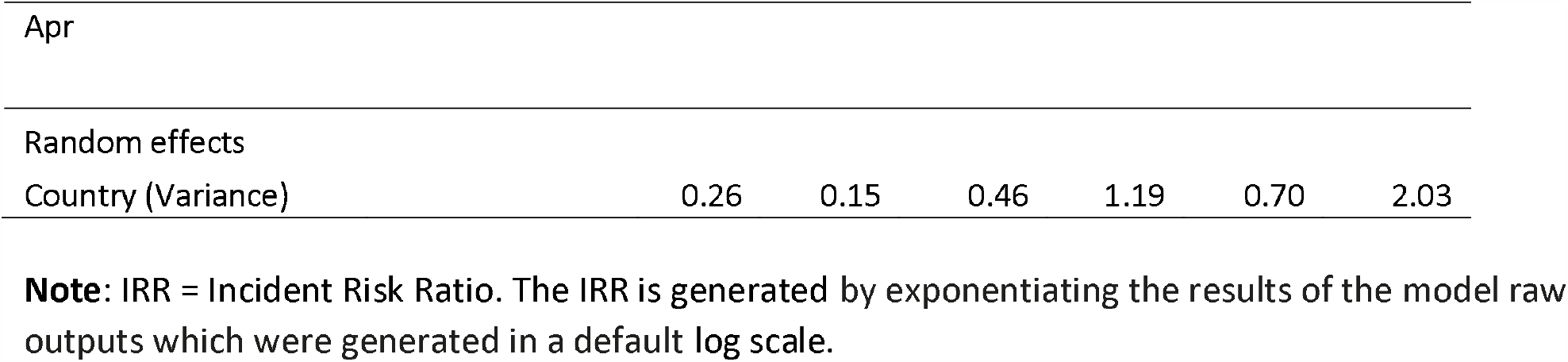
Results of hierarchical Bayesian model, effects of each intervention on case numbers and deaths

The exposure-response relationships estimated by the models are presented in Figures 1 (cases) and Figure 2 (deaths). The *X* axis represents the days since the intervention started and the *Y* axis indicates the logarithm of the risk ratio. It can be observed that mass gathering restrictions have a negative effect on the number of cases with less cases occurring as the number of days since intervention started increases. A similar effect is observed for the initial closure of business and the closure of education facilities with less cases occurring as the number of days since the intervention increases. The closure of non-essential business does not appear to have a significant effect on the number of COVID-19 cases. This is evident as the estimated relationship and its 95% credible interval stay close to zero on the Y axis. Surprisingly, stay-home measures showed a positive association with cases. This means that as the number of lock-down days increased, so did the number of cases. Negative associations were estimated for mass gatherings, initial business closure and the closure of educational facilities; while a non-significant effect was estimated for non-essential business closure. The stay-home measures showed an inverted U quadratic effect with an initial rise of cases up to day 20 of the intervention followed by a decrease in cases. These results suggest that stay at home orders may not be required to ensure outbreak control.

**Figure 1.**
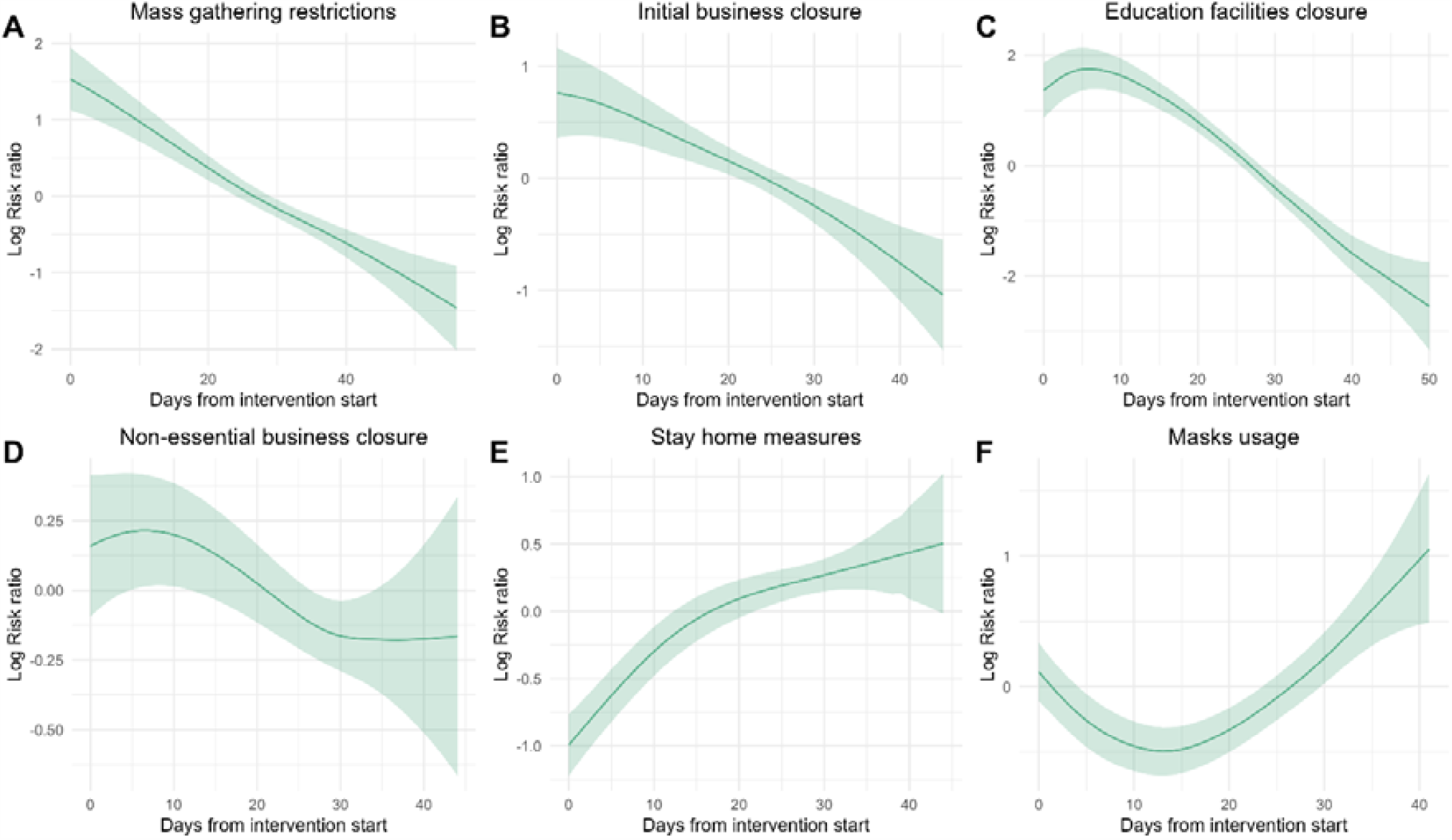
Incidence Rate Ratios (cases) following implementation of country level non-pharmaceutical control measure and daily reported COVID 19 case numbers in 30 European countries.

**Figure 2.**
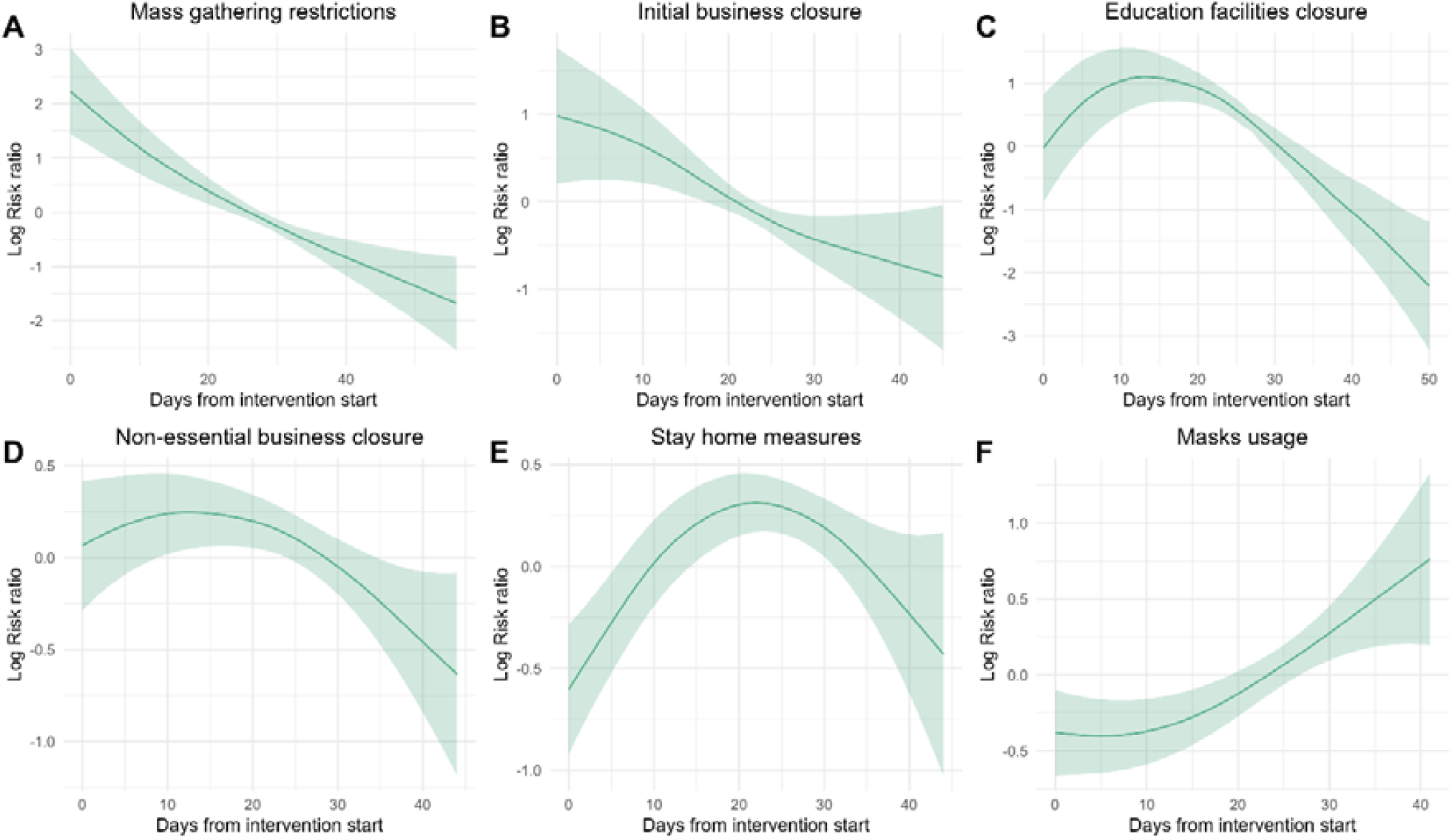
Incidence Rate Ratios (deaths) following implementation of country level non-pharmaceutical control measure and daily reported deaths from COVID-19 in 30 European countries.

Figures 3 and 4 shows the association between actual cases and deaths in each country, expressed as 7 day rolling means, and the numbers predicted by the models on cases and deaths. Although for many countries there is a reasonable correlation between the two this is not the case for all countries and particularly the smaller countries. This is most noticeable for Sweden which had lower numbers of cases and deaths than predicted, though this could be explained by partial implementation of controls and unmandated behavioural change in the population. This observation would suggest that, at least for some countries, our model does not capture all the temporally changing variables influencing the spread of the disease.

**Figure 3.**
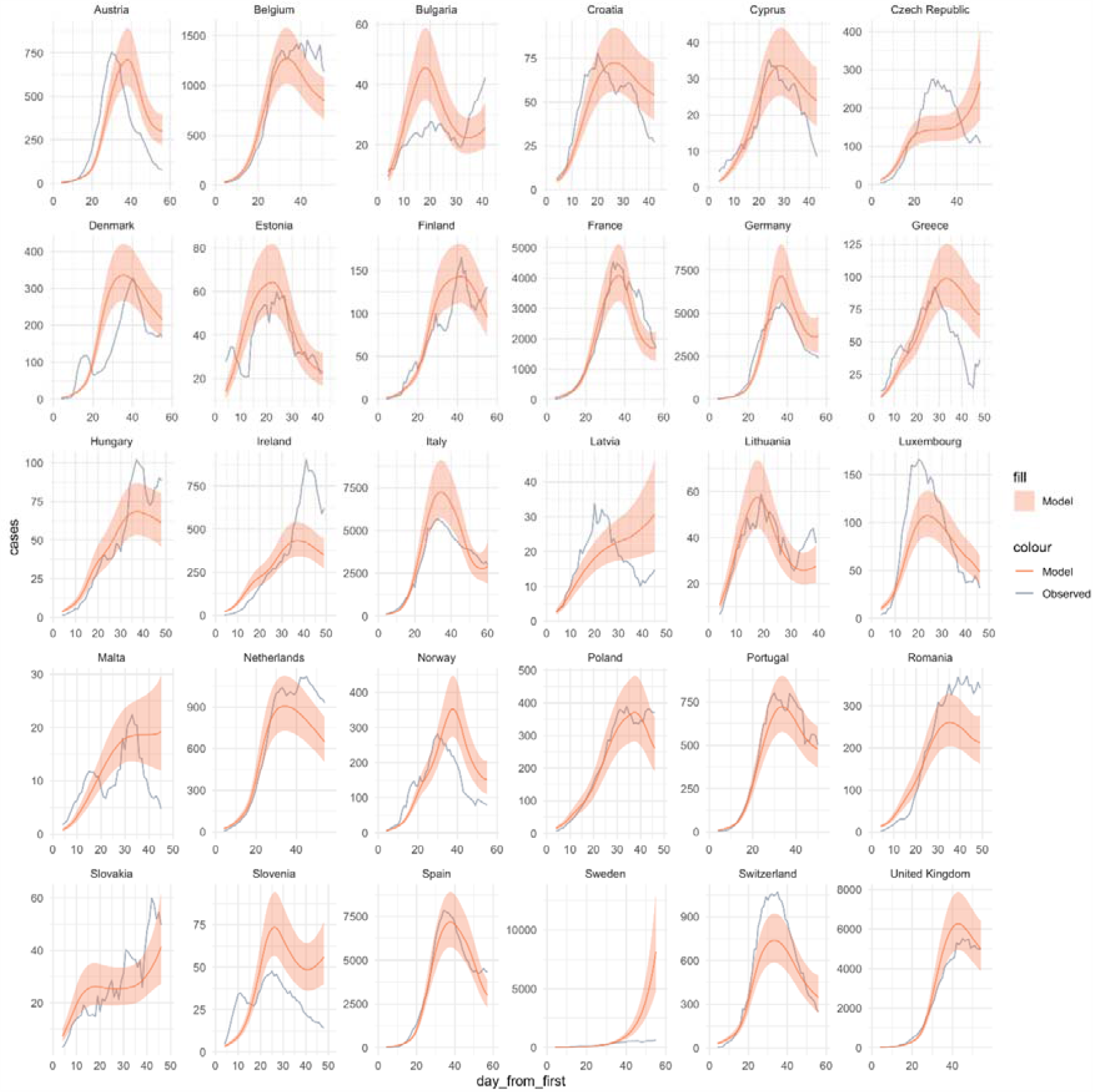
Comparison of predicted daily reports of case numbers of COVID-19 with seven day rolling average actual numbers across 30 European countries.

**Figure 4.**
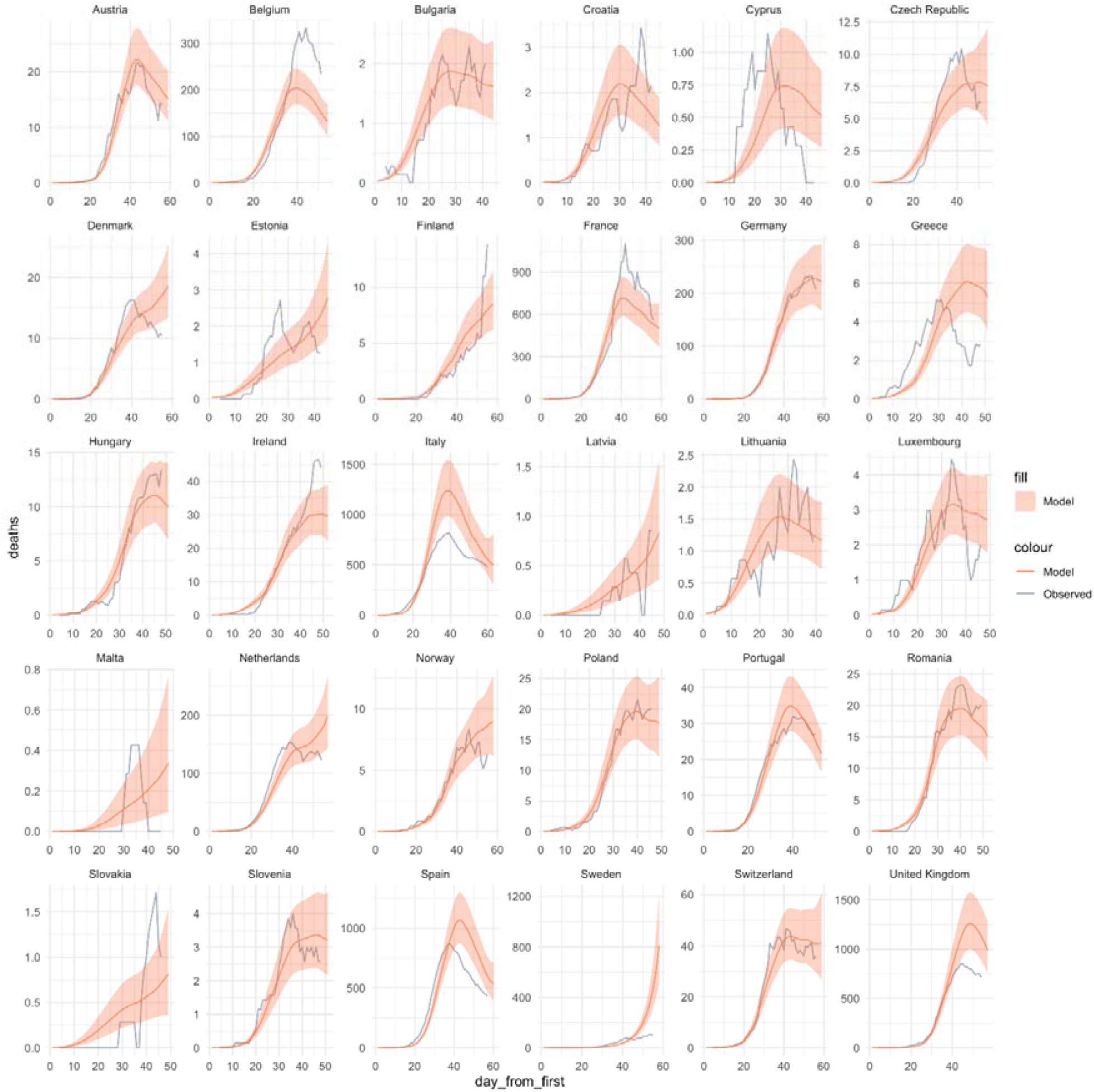
Comparison of predicted daily numbers of reports of deaths COVID-19 with seven day rolling average actual numbers across 30 European countries.

The maps of the posterior means of the country-specific risk ratios are shown in Figure 5. These maps can be interpreted as the residual risk ratio for each country after accounting for all other covariates in the model. Figure 5 also shows the country-specific posterior probability of exceeding one case or one death. The proportion of spatial variance explained by the models is 16% for the case-specific model, and 15% for the death-specific model.

**Figure 5.**
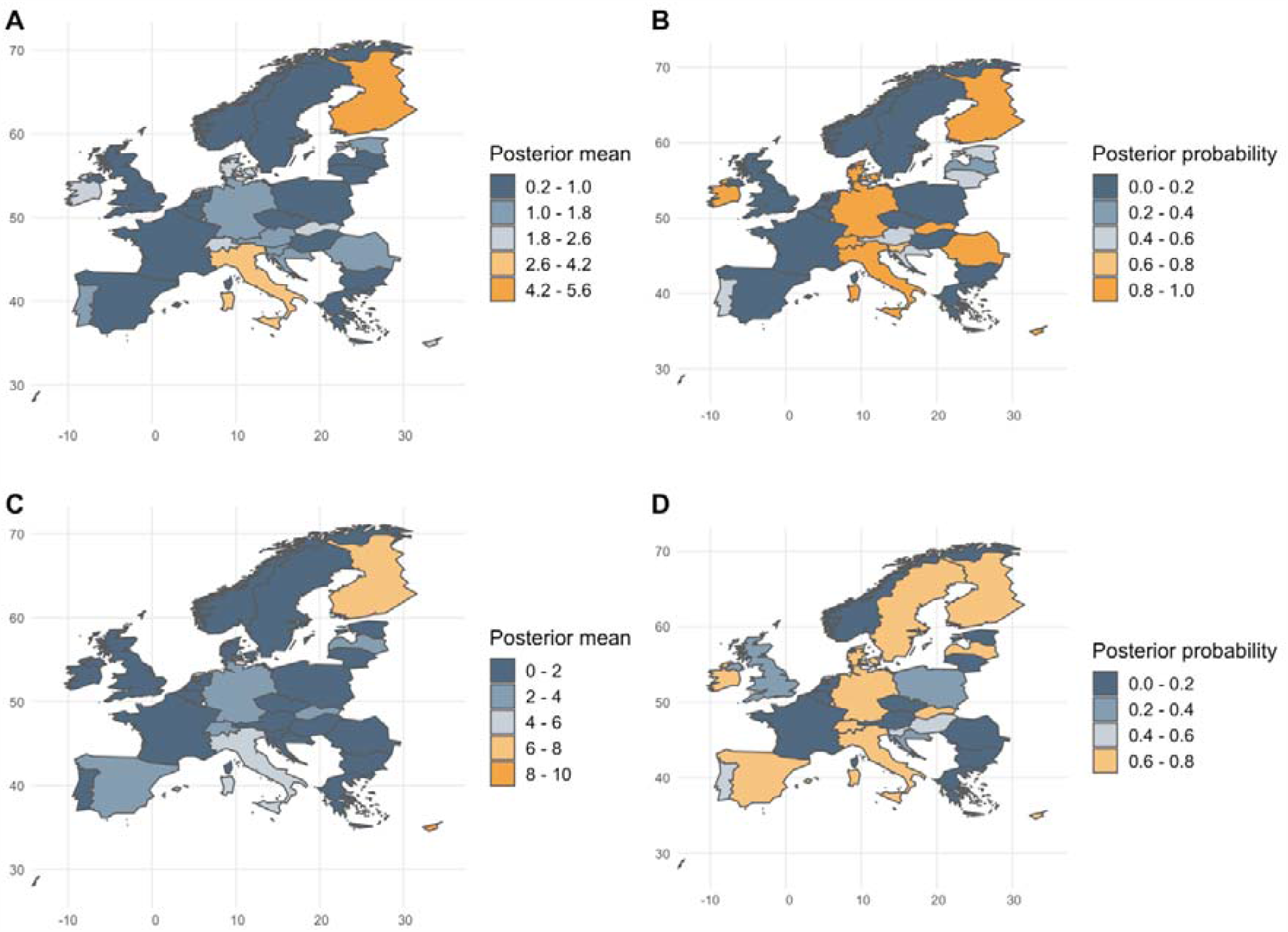
Posterior mean of the country-specific risk ratio of COVID-19 A) cases and C) deaths; and posterior probability of exceeding one COVID-19 B) case or D) death.

### Analysis 2

For confirmation and comparison, the analysis was repeated using a multilevel mixed effects model with results shown in Table 4. The conclusions of this analysis were the same as for the hierarchical probabilistic models described above. In addition, we looked at the impact of removing each intervention or all interventions on the model log likelihoods. The biggest impact came from removing educational closures from the model. The next biggest change came from removal of stay home, but this intervention was not associated with a decline in epidemic risk.

**Table 4.** Log likelihood of each model for full model compared with models excluding each of the interventions and all interventions

|  | Model | Log likelihood | Change |
| --- | --- | --- | --- |
| Cases | Full model | -9081 |  |
| Excluded | Mass gathering restrictions | -9096 | -15 |
|  | Initial business closures | -9097 | -16 |
|  | educational facilities closed | -9157 | -76 |
|  | non-essential services closed | -9085 | -4 |
|  | Stay at home advisory | -9112 | -31 |
|  | Face coverings | -9109 | -28 |
|  | All interventions | -9617 | -536 |
| Deaths | Full model | -4096 |  |
| Excluded | Mass gathering restrictions | -4101 | -5 |
|  | Initial business closures | -4109 | -13 |
|  | educational facilities closed | -4163 | -66 |
|  | non-essential services closed | -4104 | -8 |
|  | Stay at home advisory | -4113 | -17 |
|  | Face coverings | -4100 | -4 |
|  | All interventions | -4569 | -472 |

### Collinearity and sensitivity analyses

The VIF values for the predictor variables in Analysis 1 were all less that 10 (mean VIF 5.7) except for initial business closures which gave a VIF of 10.4 (Online Resource 2). Collinearity diagnostics for Analysis 2 were almost identical, in that the VIF only just exceed the 10.0 threshold and only for the initial business closures variable (Online Resource Table 3.1). The condition index exceeded 15.0 in the 9^th^ dimension (Online Resource Table 3.2) and suggested some collinearity between initial and non-essential business closure parameters. However, corresponding variance proportions in all dimensions for each control measure were well below 0.9. The smallest eigenvalue (Online Resource Table 3.3) was 0.059, which is above the suggested threshold of 0.01. These tests as a group indicate that collinearity between predictor variables did not harmfully bias the apparent separate contributions of each disease control measure (as indicated by coefficient central estimates) in our models. In addition, the standard errors of the predictors in both models were relatively small and in the sensitivity and collinearity checks, dropping each of the main predictor variables from the final equation of analysis 2 did not strongly change the coefficients and standard errors of remaining predictor variables. We conclude that there was some collinearity in our models, notably between the business closure variables, but that this was not enough to affect our conclusions.

## Discussion

We undertook a quasi-experimental study of the impact of various forms of social distancing interventions on the epidemics of COVID-19 infection in 30 different European countries. Our analyses confirm that the imposition of non-pharmaceutical control measures have been effective in controlling epidemics in each country. However, we were unable to demonstrate a strong impact from every intervention. Closure of educational facilities, banning mass gatherings and early closure of some but not necessarily all commercial businesses were all associated with reduction of the spread of infection. Widespread closure of all non-essential businesses and stay at home orders seem not to have had much additional value.

We agree with other investigators that it is difficult to separate out the effects of individual control measures when comparing between localities due to (very common) close chronological spacing of implementation (20) – but we disagree that this separation is impossible (21). We undertook sensitivity and collinearity checks and did not find that these had strongly influenced the results in each of Analysis 1 and 2. The control measures were implemented progressively in time but not in the same order in each country, or indeed implemented by all countries, so it is possible to detect separate effects.

Other studies have tried to link outbreak reduction to introduction of specific control measures using empirical data. Some of their findings concur with ours. Comparisons are especially complicated because of different categories of control measures and different outcome measures. For instance, Pan et al (22) used counts of laboratory confirmed COVID-19 infections in Wuhan, China and modelling methods to estimate the effective reproduction number (Rt) of SARS-CoV-2. They linked the Rt to many control measures: *cordons sanitaire*, traffic restriction, social distancing, home confinement, centralized quarantine, and universal symptom survey. Like us, Pan et al. divided their observations into multiple (n=5) study periods each characterised by different combinations and applications of public health measures. School closures were linked to epidemic reduction. Pan et al also concluded that the evidence was weakest for interventions implemented later (the later measures included centralised quarantine and treatment and proactive community surveillance for symptoms) because measures imposed early had already achieved much additional epidemic control. Thus, it appeared that school closures as well as stay at home orders and travel restrictions were the most effective control measures.

Zhang et al (23) looked at COVID-19 spread outside of Hubei province, China. They collected data on confirmed cases during two distinctive time periods. They estimated trends in the case counts using a Bayesian approach that included Metropolis-Hastings MCMC sampling to estimate the posterior distribution of R(t). The modelling estimated the dynamics of the net reproduction number R(t) at the provincial level. Overall, their findings suggested that strict containment measures such as travel controls, school closures, limiting movement outside the home and increased awareness within the population had contributed to interrupt local transmission of SARS-CoV-2 outside Hubei province.

COVID-19 transmission models were built by using analysis of contact surveys for periods before and after control measures were imposed in Zhang et al (24), also focusing on spread within Chinese provinces outside of Hubei. The outcome was estimates of R_0_. These authors performed generalised linear mixed model regression to account for clustering and potential correlations (multiple contacts such as those in same household exposed to the same index case). The models adjusted for age, gender, type of contact and travel history of individuals. The authors concluded that school closures could have large impacts on disease dynamics although school closures in isolation were insufficient to prevent or stop a COVID-19 outbreak. Because most early transmissions before control measures started occurred between members of the same household, stay at home measures were not as important for disease control, but in contrast, travel restrictions and prohibition of large gatherings did reduce transmissions.

It seems likely that many possible combinations of social distancing measures can be effective, although we only tried to analyse the impacts seen in the European context. The order of measures may have influenced both our analysis and those of other studies (described above) that also tried to assess which control measures were most effective in epidemic reduction. Measures imposed later may seem less effective simply because of their place in the order that they happened (additional benefits were small after other measures were put in place). Our analyses indicated that school closures and stopping mass gatherings were most effective, but we acknowledge that this had to be at least somewhat dependent on the order of interventions in Europe. Different measures reinforced and enabled each other: for instance, there was little incentive to leave home if schools and businesses are already closed and weather was inclement (as it often is in early spring in Europe, when most national lockdowns started). Business and school closures usually preceded stay at home measures in Europe, so it may not have been possible for data on stay at home orders to be linked to large additional effects. This potential ordering problem is at least somewhat mitigated for by our use of individual lag measures (in timing) from when each intervention was effected. It is also worth noting that outside of institutional and crowded settings, there is evidence that much if not most COVID-19 transmission is within households (25); stay at home orders intensify contact within households which would be expected to increase household transmission. It could be therefore not surprising that stay at home measures on their own are not very effective outbreak control measures and may not generate large additional benefits.

Spatiotemporal hierarchical models, like the one we presented, have the advantage of being able to explicitly quantify the probability that an epidemic may or may not occur at a specific time or location. Public health officials may be more inclined to deploy interventions if the probability of an epidemic exceeds tolerable thresholds. If such thresholds can be clearly measured, ideally, public health decision-makers could agree on the specific epidemic thresholds (i.e. the incidence above which the disease requires imposition of control measures) to make model predictions useful for decision support.

Whether or not school closures are likely to have been important in controlling the spread of epidemic disease is an issue of some debate in both the scientific and lay media. There has been uncertainty about how beneficial the closing of educational establishments can be on respiratory disease transmission (26-28). The value of school closures is particularly uncertain for COVID-19 given the observation that children have only mild or no symptoms (29). Decline in the infectiousness of the SARS-1 outbreak in Hong Kong in 2003 was also a time when many interventions were implemented, including school closures (30), making it hard to disentangle contributions of each individual measure. In the current pandemic, Hong Kong managed to substantially reduce the transmissibility of COVID-19 fairly early in the outbreak by a limited number of interventions one of which was keeping schools closed (31). However, there were other substantial behavioural change in the population at the time coincident with these interventions. Viner and colleagues (26) state “Data from the SARS outbreak in mainland China, Hong Kong, and Singapore suggest that school closures did not contribute to the control of the epidemic”. However, this is not a valid argument against school closures as peak shedding of virus in SARS-CoV was around day 10 whereas peak shedding of SARS-CoV-2 is much earlier and possibly before symptoms develop (32-34). In contrast to COVID-19, SARS was known to be primarily infectious after onset of symptoms when most cases would have been hospitalised or at least quarantined. Throat swabs from children have shown similar viral load to those in adults, yet contact tracing studies tend to find few incidents where transmission occurred from children to adults (35, 36). We cannot resolve the lack of consensus in these lines of evidence, about how likely children are to pass SARS-COV-2 to adults. What our study also does not do is identify which level of school closure has the most benefit whether it is primary, junior, senior school or even higher education. This will need to be the focus of further research. Note also that the results presented here are based on total closure rather than schools operating completely as normal (with no physical distancing). The utility of social distancing may be much more important between adult staff rather than between child pupils; given the body of evidence that suggest little SARS-CoV-2 transmission has occurred between children while there is ample evidence of transmission between adults (36, 37). Hence, transmission may be significantly reduced if adult staff within schools observe social distancing from other adults even if pupils do not physical distance from each other. It is also possible that reduced school opening such as three-day weekends could have worthwhile impacts on the spread of infection (38).

The second greatest impact on the epidemiology of the European COVID-19 was from banning mass gatherings (which could be of any size), both public and private gatherings. A 2018 review of the impact of mass gatherings on outbreaks of respiratory infectious disease (39) found that most evidence was linked to the Islamic Hajj pilgrimage, where most infections were respiratory, mainly rhinovirus, human coronaviruses and influenza A virus. The evidence for respiratory disease outbreaks arising from other mass gatherings such as music festivals or sporting events is less established, but not absent. Several outbreaks of respiratory infectious disease have been linked to open air festivals and other music festivals (39, 40). For instance, during the 2009 influenza season pandemic influenza A(H1N1)pdm09 outbreaks were recorded at three of Europe’s six largest music festivals, while some 40% of pandemic flu cases that season in Serbia were linked with the Exit music festival.

The types of business closures are interesting. We established that there was weak collinearity between the two types of business closures in the models. However, the stronger association was with the initial business closures. Given that those initial closures were mostly directed at business where people congregate and have a purpose of facilitating socialising (i.e. the hospitality industry), this would suggest that control measures among these businesses are where the most impact may be had. Although outbreaks of food poisoning are frequently linked with venues where food is consumed, outbreaks of respiratory infections are much more rarely so. One exception was an outbreak of SARS at a restaurant where live palm civets were caged close to customer seating (41). The link with COVID-19 is probably much less about food and beverage consumption, and simply about time people spend in close proximity to each other.

### Limitations

Although our study suggests that closures of educational interventions and banning mass gatherings are the most important measures, this is caveated with several observations. Many interventions were implemented in different ways and at different points in the local epidemic. For example, in accordance with the IHME assignment, we treated Sweden as a country without school closures because schools for persons under 16 stayed open, although upper secondary and tertiary education facilities were actually shut in Sweden from late March 2020 (42). Similarly, the exact timing of restrictions being introduced varied over time in Italy, Spain and between individual federal states in Germany (43). Which types of work places could stay open varied; while the acceptable reasons for being outdoors also varied between countries. Stay at home orders in some countries was an advisory but not enforced whilst elsewhere stay-home orders were enforced by police with penalties. In some countries, children could go outside and outdoor exercise was permitted whilst in others either or both might be banned. In some countries, severe travel restrictions were a separate intervention whilst in others travel bans were a consequence of a stay at home order and could not be identified separately. Because of this variety in how interventions were implemented and described, the results for the potential of stay at home advisories especially may be under-estimated. All models are simplifications of the complex nature of reality; our modelling was unable capture many subtle variations in how control measures were implemented. We acknowledge that lack of direct observation of these variations may have biased our results.

## Conclusion

Relaxing stay-at-home orders and allowing reopening of non-essential businesses appeared to be the lowest risk measures to relax as part of plans to carefully lift COVID-19 lockdown measures. There is still even now relatively little unclear empirical evidence on the relative value of different interventions. And yet, the reasons to implement minimal control measures are compelling, given the social and economic harm linked to tight control measures. Hence, whilst we need to be cautious about using preliminary results, public health officials will have to use evidence as it emerges rather than expect to wait for a final full view to decide what might be (was) the best control strategy. Careful monitoring of how relaxation of each control measure affects transmissibility of COVID-19 is required and will help to minimise the inevitably imperfect results.

## Data Availability

Data in public domain. Data and model code can be shared upon request to authors.

## REFERENCES

1. Jacobs LA. Rights and quarantine during the SARS global health crisis: differentiated legal consciousness in Hong Kong, Shanghai, and Toronto. Law & Society Review. 2007;41(3):511–52.

2. Johnson HC, Gossner CM, Colzani E, et al. Potential scenarios for the progression of a COVID-19 epidemic in the European Union and the European Economic Area, March 2020. Eurosurveillance. 2020;25(9):2000202.

3. Brooks SK, Webster RK, Smith LE, et al. The psychological impact of quarantine and how to reduce it: Rapid review of the evidence. The Lancet. 2020.

4. Office for Budget Responsibility. Coronavirus reference scenario. 2020. https://obr.uk/coronavirus-reference-scenario/.

5. Massaro E, Ganin A, Perra N, Linkov I, Vespignani A. Resilience management during large-scale epidemic outbreaks. Scientific Reports. 2018;8(1):1–9.

6. Adam D. Special report: The simulations driving the world’s response to COVID-19. Nature. 2020.

7. Sridhar D, Majumder MS. Modelling the pandemic. British Medical Journal 2020;369:m1567. doi:10.1136/bmj.m1567

8. Borenstein S, Johnson CK. Modeling coronavirus: ‘Uncertainty is the only certainty’. Associated Press,. 2020 7 April.

9. Andrews C. Predicting the pandemic: mathematical modelling tackles Covid-19. Engineering & Technology. 2020 10 April.

10. McCoy D. Faith in coronavirus modelling is no substitute for sound political judgment. The Guardian. 2020 10 April.

11. Rosenbaum PR. How to see more in observational studies: Some new quasi-experimental devices. Annual Review of Statistics and Its Application. 2015;2:21–48.

12. Obaromi D. Spatial Modelling of Some Conditional Autoregressive Priors in A Disease Mapping Model: the Bayesian Approach. Biomedical Journal of Scientific & Technical Research. 2019;14(3). doi:10.26717.BJSTR.2019.14.002555

13. Chatterjee S, Hadi AS. Regression analysis by example. 4th ed: John Wiley & Sons; 2015.

14. Regorz A. How to interpret a Collinearity Diagnostics table in SPSS. 2020. http://www.regorz-statistik.de/en/collinearity_diagnostics_table_SPSS.html. Accessed Jun 24 2020.

15. Bi Q, Wu Y, Mei S, et al. Epidemiology and transmission of COVID-19 in 391 cases and 1286 of their close contacts in Shenzhen, China: a retrospective cohort study. The Lancet Infectious Diseases. 2020.

16. Backer JA, Klinkenberg D, Wallinga J. Incubation period of 2019 novel coronavirus (2019-nCoV) infections among travellers from Wuhan, China, 20–28 January 2020. Eurosurveillance. 2020;25(5):2000062.

17. Lauer SA, Grantz KH, Bi Q, et al. The incubation period of coronavirus disease 2019 (COVID-19) from publicly reported confirmed cases: estimation and application. Annals of Internal Medicine. 2020;172(9):577–82.

18. Wright O. People told to wait ten days for coronavirus test results. The Times. 2020 May 7.

19. Zhou F, Yu T, Du R, et al. Clinical course and risk factors for mortality of adult inpatients with COVID-19 in Wuhan, China: a retrospective cohort study. The Lancet. 2020.

20. Hsiang S, Allen D, Annan-Phan S, et al. The effect of large-scale anti-contagion policies on the coronavirus (covid-19) pandemic. MedRxiv. 2020.

21. Flaxman S, Mishra S, Gandy A, et al. Estimating the number of infections and the impact of non-pharmaceutical interventions on COVID-19 in 11 European countries. Nature. 2020.

22. Pan A, Liu L, Wang C, et al. Association of public health interventions with the epidemiology of the COVID-19 outbreak in Wuhan, China. Journal of the American Medical Association. 2020.

23. Zhang J, Litvinova M, Wang W, et al. Evolving epidemiology and transmission dynamics of coronavirus disease 2019 outside Hubei province, China: a descriptive and modelling study. The Lancet Infectious Diseases. 2020.

24. Zhang J, Litvinova M, Liang Y, et al. Changes in contact patterns shape the dynamics of the COVID-19 outbreak in China. Science. 2020.

25. Koh WC, Naing L, Rosledzana MA, et al. What do we know about SARS-CoV-2 transmission? A systematic review and meta-analysis of the secondary attack rate, serial interval, and asymptomatic infection. medRxiv. 2020.

26. Viner RM, Russell SJ, Croker H, et al. School closure and management practices during coronavirus outbreaks including COVID-19: A rapid systematic review. The Lancet Child & Adolescent Health. 2020.

27. Cowling BJ, Lau EH, Lam CL, et al. Effects of school closures, 2008 winter influenza season, Hong Kong. Emerging Infectious Diseases. 2008;14(10):1660.

28. Hens N, Ayele GM, Goeyvaerts N, et al. Estimating the impact of school closure on social mixing behaviour and the transmission of close contact infections in eight European countries. BMC Infectious Diseases. 2009;9(1):187.

29. Shen K, Yang Y, Wang T, et al. Diagnosis, treatment, and prevention of 2019 novel coronavirus infection in children: experts’ consensus statement. World Journal of Pediatrics. 2020:1–9.

30. Riley S, Fraser C, Donnelly CA, et al. Transmission dynamics of the etiological agent of SARS in Hong Kong: impact of public health interventions. Science. 2003;300(5627):1961–6.

31. Cowling BJ, Ali ST, Ng TW, et al. Impact assessment of non-pharmaceutical interventions against coronavirus disease 2019 and influenza in Hong Kong: an observational study. The Lancet Public Health. 2020.

32. Pan Y, Zhang D, Yang P, Poon LL, Wang Q. Viral load of SARS-CoV-2 in clinical samples. The Lancet Infectious Diseases. 2020;20(4):411–2.

33. Peiris JSM, Chu C-M, Cheng VC-C, et al. Clinical progression and viral load in a community outbreak of coronavirus-associated SARS pneumonia: a prospective study. The Lancet. 2003;361(9371):1767–72.

34. Wei WE, Li Z, Chiew CJ, Yong SE, Toh MP, Lee VJ. Presymptomatic Transmission of SARS-CoV-2—Singapore, January 23–March 16, 2020. Morbidity and Mortality Weekly Report. 2020;69(14):411.

35. Boast A, Munro A H. G. DFTB COVID-19 Evidence Review: UK Royal College of Paediatrics and Child Health 2020 22 April.

36. Viner RM, Mytton OT, Bonell C, et al. Susceptibility to and transmission of COVID-19 amongst children and adolescents compared with adults: a systematic review and meta-analysis. medRxiv. 2020:27.

37. Victoria State government. Physical distancing, health and hygiene. 2020. https://education.vic.gov.au/parents/Pages/Physical-distancing,-health-and-hygiene.aspx. Accessed June 20 2020.

38. Cooley PC, Bartsch SM, Brown ST, Wheaton WD, Wagener DK, Lee BY. Weekends as social distancing and their effect on the spread of influenza. Computational and Mathematical Organization Theory. 2016;22(1):71–87.

39. Hoang V-T, Gautret P. Infectious diseases and mass gatherings. Current Infectious Disease Reports. 2018;20(11):44.

40. Botelho-Nevers E, Gautret P. Outbreaks associated to large open air festivals, including music festivals, 1980 to 2012. Eurosurveillance. 2013;18(11):20426.

41. Wang M, Yan M, Xu H, et al. SARS-CoV infection in a restaurant from palm civet. Emerging Infectious Diseases. 2005;11(12):1860.

42. Johnson S. Swedish parents fret as schools stay open amid European virus shutdown. Reuters. 27 March 2020.

43. Anonymous. How are Germany’s 16 states dealing with the coronavirus pandemic? The Local DE. 19 March 2020.

44. Watanabe S. Asymptotic equivalence of Bayes cross validation and widely applicable information criterion in singular learning theory. Journal of Machine Learning Research. 2010;11(Dec):3571–94.

45. Pettit L. The conditional predictive ordinate for the normal distribution. Journal of the Royal Statistical Society: Series B (Methodological). 1990;52(1):175–84.

